# Contamination of personal protective equipment during COVID-19 autopsies

**DOI:** 10.1101/2021.07.12.21260357

**Authors:** Johanna M. Brandner, Peter Boor, Lukas Borcherding, Carolin Edler, Sven Gerber, Axel Heinemann, Julia Hilsenbeck, Atsuko Kasajima, Larissa Lohner, Bruno Märkl, Jessica Pablik, Ann Sophie Schröder, Linna Sommer, Julia Slotta-Huspenina, Jan-Peter Sperhake, Saskia von Stillfried, Sebastian Dintner

**Author notes:** **Corresponding author:** Prof. Bruno Märkl, General Pathology and Molecular Diagnostics, Medical Faculty, University Augsburg, Germany, Full postal address: Stenglinstrasse 2, 86156 Augsburg. **Competing Interests Statement:** The authors have nothing to disclose.

## Abstract

Confronted with an emerging infectious disease, the medical community faced relevant concerns regarding the performance of autopsies of COVID-19 deceased at the beginning of the pandemic. This attitude has changed, and autopsies are now recognized as indispensable tools for elucidating COVID-19; despite this, the true risk of infection for autopsy staff is still debated. To elucidate the rate of SARS-CoV-2 contamination in personal protective equipment (PPE), swabs were taken at nine locations of the PPE of one physician and an assistant each from 11 full autopsies performed at four different centers. Further samples were obtained for three minimally invasive autopsies (MIA) conducted at a fifth center. Lung/bronchus swabs of the deceased served as positive controls. SARS-CoV-2 RNA was detected by RT-qPCR. In 9/11 full autopsies PPE samples were tested RNA positive with PCR, in total 21% of all PPE samples taken. The main contaminated parts of the PPE were the gloves (64% positive), the aprons (50% positive), and the upper sides of shoes (36% positive) while for example the fronts of safety goggles were only positive in 4.5% of the samples and all face masks were negative. In MIA, viral RNA was observed in one sample from a glove, but not in other swabs. Infectious virus isolation in cell culture was performed in RNA positive swabs from full autopsies. Of all RNA positive PPE samples, 21% of the glove samples were positive for infectious virus taken in 3/11 full autopsies. In conclusion, in >80% of autopsies, PPE was contaminated with viral RNA. In >25% of autopsies, PPE was found to be even contaminated with infectious virus, signifying a potential risk of infection among autopsy staff. Adequate PPE and hygiene measures, including appropriate waste deposition, are therefore mandatory to enable safe work environment.

## Introduction

The results obtained through autopsies of the Severe acute respiratory syndrome Coronavirus-2 (SARS-CoV-2)-infected deceased are of crucial importance for understanding Coronavirus disease 2019 (COVID-19). Viral pneumonia with diffuse alveolar damage (DAD) is the most frequent cause of death in fatal cases of COVID-19. In addition to the dramatic changes in the lungs, the affection of multiple other organs is currently interpreted mainly as a systemic inflammatory reaction. Several authors have described endothelial impairment with consecutive activation of the coagulation system (1, 2).

However, concerns about the safety of autopsy staff hampered the autopsy activities of surgical, forensic and neuro-pathologists. Since the beginning of the pandemic, several reports and guidelines from different authors and organizations have been published (3, 4) concerning this topic.

The environmental viability of the virus has been investigated in experimental conditions and real-world settings of the domestic or clinical surroundings of SARS-CoV-2-positive persons in a few studies which were recently summarized by Meyerowitz et al. (5). The presence of viable virus has been identified for up to three hours in aerosols and 72 hours on surfaces. Half-lives were calculated to be up to six hours (6).

In Germany, the Robert Koch Institute has recommended compliance with protection level 3, which requires wearing appropriate protective equipment (surgical hood cap, eye/face-protection with fully protective safety goggles or visors, filtering face piece (FFP) 2/3 masks, long-sleeved and impermeable protective clothing, waterproof apron, additional forearm protection, a second layer of latex/nitrile gloves with long cuffs, and appropriate shoes), when handling COVID-19 deceased (7).

Only a few reports addressing topics related to the infectiousness of dead bodies of the SARS-CoV-2-infected deceased and the risk for autopsy staff have been published. All authors report detection of the virus with reverse transcription quantitative PCR (RT-qPCR) in swabs taken from the airways at different time intervals after death (8-11). Schroeder et al. detected viral RNA on various body surfaces of the deceased, as well as body bags; however, no viable viruses were detected (12). Viral RNA detection on the surfaces of the autopsy tables and autopsy room walls as well as face shields was reported by Pomara et al. (11).

The aim of this study is to evaluate the extent of viral RNA contamination of the personal protective equipment (PPE) of autopsy staff during autopsies of the COVID-19 deceased. Special focus was placed on the infectivity of the samples with positive SARS-CoV-2 RNA detection.

## Materials and methods

### Participating centers and case collection

This study was conducted within the German framework of the DEFEAT PANDEMIcs initiative, which aims, among other objectives, to develop an operational and organizational basis for autopsy programs at the national level for pandemic preparedness. Four clinical pathology departments (Aachen [AA], Augsburg [AU], Dresden [DR], and Munich [MU]) and one department of legal medicine (Hamburg [HH]) participated in this study between January and May 2021. Four centers (AA, AU, DR, and HH) performed complete autopsies with the opening of all body cavities, including the skulls. For the latter, only oscillating saws with vacuum were used for safety reasons. Minimally invasive autopsies (MIA) with ultrasound-conducted biopsies were performed at MU. Written consent was obtained from the next of kin to perform the autopsies. The inclusion criterion for decedents was confirmed diagnosis of SARS-CoV-2 infection, as evidenced by PCR test of the nasopharyngeal swab during the hospital stay and by either rapid PCR or antigen testing during the full autopsies. Three autopsies each from AU, DR, HH, and MU were included, and two cases were contributed by AA. All autopsy rooms were maintained at negative pressure, with a minimum of 10 air changes per hour.

### Swabs – Specimen collection and locations

Commercially available swab sets were used (COPAN eSwab B 80482CE, Mast Group, Reinfeld, Germany). Before swabbing, the tips were moistened with the transport medium, and then the PPE surfaces were thoroughly swabbed in a meandering manner for at least 15 seconds. Finally, the tips were placed in the transport container. The swab locations are shown in Fig 1. Two swabs per location were taken next to each other, one for RT-qPCR testing, one for testing of virus infectivity (virus isolation). Samples for RT-qPCR testing were stored in refrigerators at 4°C, while swabs intended for eventual isolation of infectious virus were frozen at -80°C. PCR testing of all samples was performed at AU. For samples from full autopsies that showed positive results in the PCR testing, the corresponding samples collected for viral isolation were sent to HH, where another PCR testing and virus isolation were performed.

**Fig. 1.**
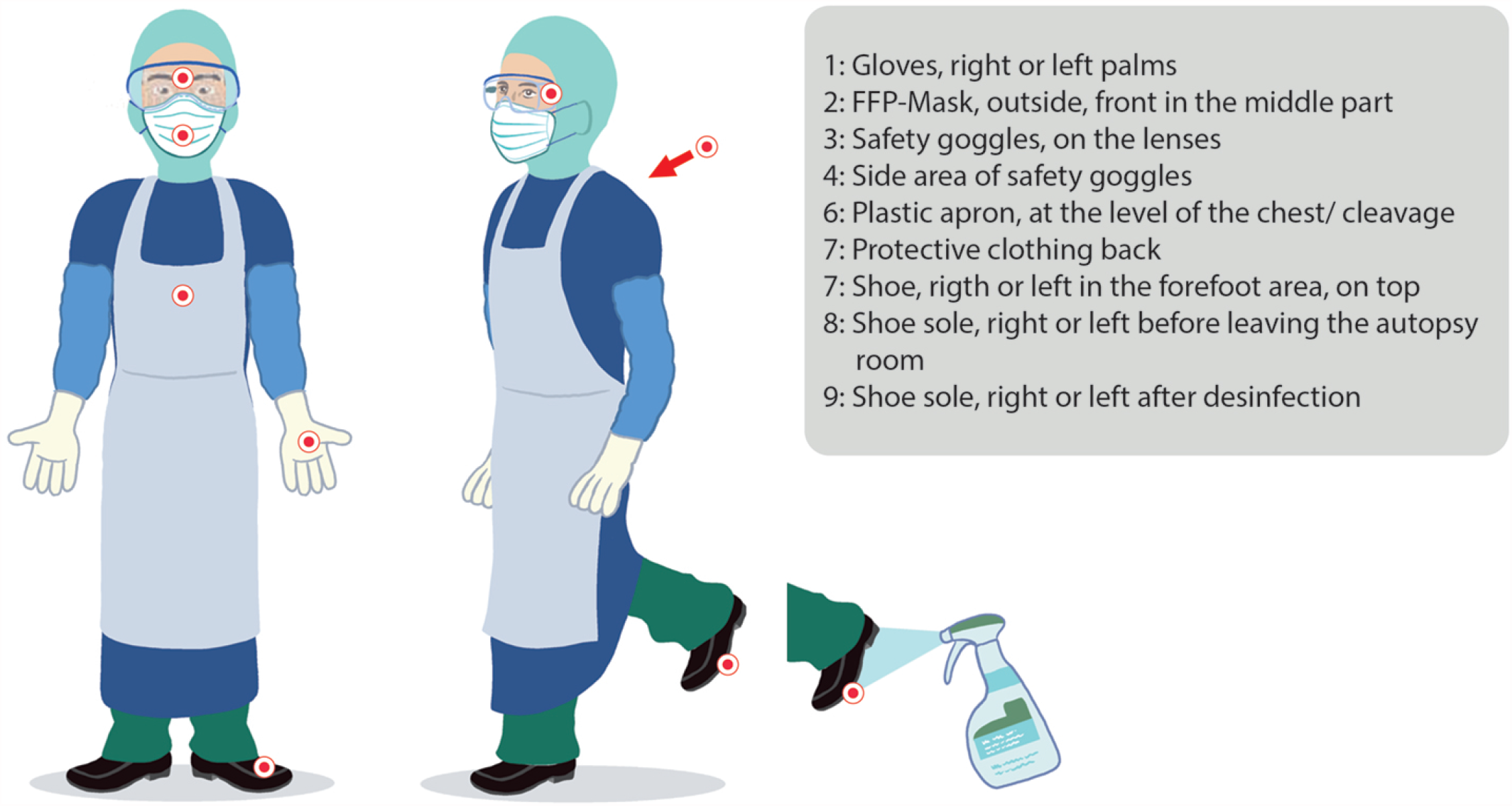
Schematic representation of the swab collection localizations.

In all full autopsies, swabs from the PPE of the autopsy-conducting physician and one autopsy assistant were performed after finishing the autopsy. In MIAs, only the team member with the closest contact to the body of the deceased – the ultrasound-guiding physician – was evaluated.

To generate reference samples (positive controls), swabs from the plane-cut surface of the lungs and, in one case, the bronchus (case AU2) or from the lung biopsies (MIA, MU) were collected during each autopsy.

### RT-qPCR

All samples were processed and analyzed primarily at the Institute of Pathology and Molecular Diagnostics at the Medical Center in Augsburg. The method has been described in the literature (13). In brief, RNA was extracted using the Promega Maxwell® 16 MDx system and the Promega Maxwell 16 LEV RNA FFPE Purification Kit (AS1260, Promega Corporation, Madison, WI, USA). Quantitative real-time PCR for SARS-CoV-2 was performed on the extracts with one-step multiplex RT-qPCR targeting the SARS-CoV-2 ORF1ab, N Protein, and S Protein using the TaqPath COVID-19 CE-IVD RT-PCR Kit (A48067, Thermo Fisher, Pleasanton, TX, USA). The RT-qPCR was conducted using the QuantStudio 5 Dx real-time PCR Instrument, and the data were analyzed and interpreted using QuantStudio™ design and analysis software (v.1.2x, Thermo Fisher, Carlsbad, CA, USA). Results with two or more positive targets were considered as valid. A singular failure of the curve for the S protein was used as indirect evidence for the presence of a virus variant. A verification was carried out by comparing it with the results of the mutational diagnostics during the clinical stay.

### Cell culture and virus isolation

Duplicate swabs stored at -80°C for isolation of infectious virus from locations with RT-qPCR positive swabs (as determined in AU) were transferred to the biosafety level (BSL)-3 laboratory at the Institute of Medical Microbiology, Virology, and Hygiene at the University Medical Center Hamburg-Eppendorf. For control of RNA integrity, confirmatory RT-qPCR of the samples was performed as described (14). Vero E6 cells were maintained and cultivated under standard conditions (15). For virus isolation, 500 μl of swab medium was used, and infection was performed as described (12, 15). Supernatants were harvested at 72 hours post-infection, and virus growth was analyzed, as described before (16). Virus isolation experiments were restricted to samples from full autopsies.

### Statistics

To compare data measured by order or rank, the Mann-Whitney-U-Test and the One Way Repeated Measures ANOVA test were used. The Spearman’s rank order correlation was applied to calculate correlations between ranked data. Depending on the proportion numbers, tabulated nominal data were compared using either the chi-square test or Fisher’s exact test. A p-value < 0.05 was considered significant. All calculations were performed using the statistics package Sigmaplot 13.0 (Systat, San Jose, CA, USA).

## Results

### Case collection

The case characteristics are given in Table 1. Fourteen autopsies were included, of which three were conducted as MIA. The median age of the deceased was 71 years (range: 52 – 91 years), with a male to female ratio of 1.8: 1. The postmortem interval (PMI) had a broad range (15 to 144 hours, median 55 hours). The median period from the first positive SARS-CoV-2 RT-PCR test to death was 10.5 days (range: 0 – 51 days).

**Table.** Demographic and autoptic data of all cases. Rf = reference organ (positive control); Gl = Gloves; n.a. = no data available; n.i. = no information concerning the viral lineage available; * evaluated during RT-qPCR – loss of the S-curve as hint for variant of concern; ** no testing prior to death #:positive tested in confirmatory RT-qPCR in HH.

|  | Autopsy/<br>Case-No | Gender | Age [5<br>year<br>intervals] | BMI<br>[kg/m<sup>2</sup>] | Time<br>between<br>1st positive<br>test and<br>death<br>[days] | Time<br>between<br>death and<br>autopsy<br>[hours] | Location of<br>death | Cause of death | Ct-value<br>reference<br>sample at<br>autopsy [N-gene] | Infectious<br>Virus | Variant of<br>concern* |
| --- | --- | --- | --- | --- | --- | --- | --- | --- | --- | --- | --- |
| Full autopsies | 1 (AA-1) | male | 70-74 | 22 | 28 | 38 | hospital | DAD - COVID-19 | 24 | no | no |
|  | 2 (AA-2) | female | 85-89 | 23 | 25 | 64 | hospital | DAD - COVID-19 | 28 | no | no |
|  | 3 (AU-1) | male | 85-89 | 25 | 8 | 15 | hospital | DAD - COVID-19 | 15 | Rf /GI | no |
|  | 4 (AU-2) | female | 75-79 | 36 | 13 | 40 | hospital | DAD - COVID-19 | 28 | no | no |
|  | 5 (AU-3) | female | 90-94 | 16 | 19 | 36 | hospital | DAD - COVID-19 | 17 | Rf | no |
|  | 6 (DR-1) | male | 60-64 | 50 | 7 | 96 | hospital | DAD - COVID-19 | 16 | no | no |
|  | 7 (DR-2) | male | 80-84 | 36 | 10 | 120 | hospital | DAD - COVID-19 | 16 | Rf/GI | no |
|  | 8 (DR-3) | male | 65-69 | 35 | 3 | 72 | hospital | combined hepatic and cardial failure | 14 | GI | yes |
|  | 9 (HH-1) | male | 65-69 | 68 | 2 | 48 | outpatient | Systemic inflammatory due to<br>pyelonephritis and pneumonia | 15 | no | no |
|  | 10 (HH-2) | female | 50-54 | 53 | n.a. | 144 | hospital | DAD - COVID-19 | 32<sup>#</sup> | no | no |
|  | 11 (HH-3) | female | 70-74 | 30 | 0** | 144 | outpatient | DAD - COVID-19 | 22 | Rf | yes |
| MIA | MU-1 | male | 60-64 | 18 | 11 | 21 | hospital | n.a. | 20 | / | yes |
|  | MU-2 | male | 50-54 | 47 | n.a. | 62 | hospital | acute respiratory failure due to<br>pneumothorax | negative | / | n.i. |
|  | MU-3 | male | 70-74 | 23 | 51 | 22 | hospital | n.a. | negative | / | n.i. |

### RT-qPCR results of swabs in full autopsies

In total, 209 swabs were performed for 11 full autopsies for RT-qPCR testing. Eleven of these samples were taken from the lungs/bronchus of the dead body to serve as a reference (positive control) for each case. All lung/bronchus swabs (11/11) were positive.

The remaining 198 swabs were collected from nine locations on the PPE of one physician and one assistant per autopsy. Of these, 41 (21%) were SARS-CoV-2 RNA-positive, 24 from physicians and 17 from assistants (Fig. 2A). In only two autopsies (2/11, 18%) all PPE swabs were SARS-CoV-2 RNA negative, (Fig. 2A), while in 9/11 autopsies (82%) RNA contamination was detected with the number of SARS-CoV-2 RNA-positive PPE swabs per autopsy ranging from 3 to 7 (median 4, Fig. 2A). No correlation was observed between the PMI and the number of RNA positive PPE swabs (p = 0.503; Fig. 2A). In addition, the total number of positive swabs per autopsy did not correlate with the cycle threshold (Ct) values of the lungs/bronchus swabs (R = -0.51; p = 0.126) (Table 1).

**Fig. 2.**
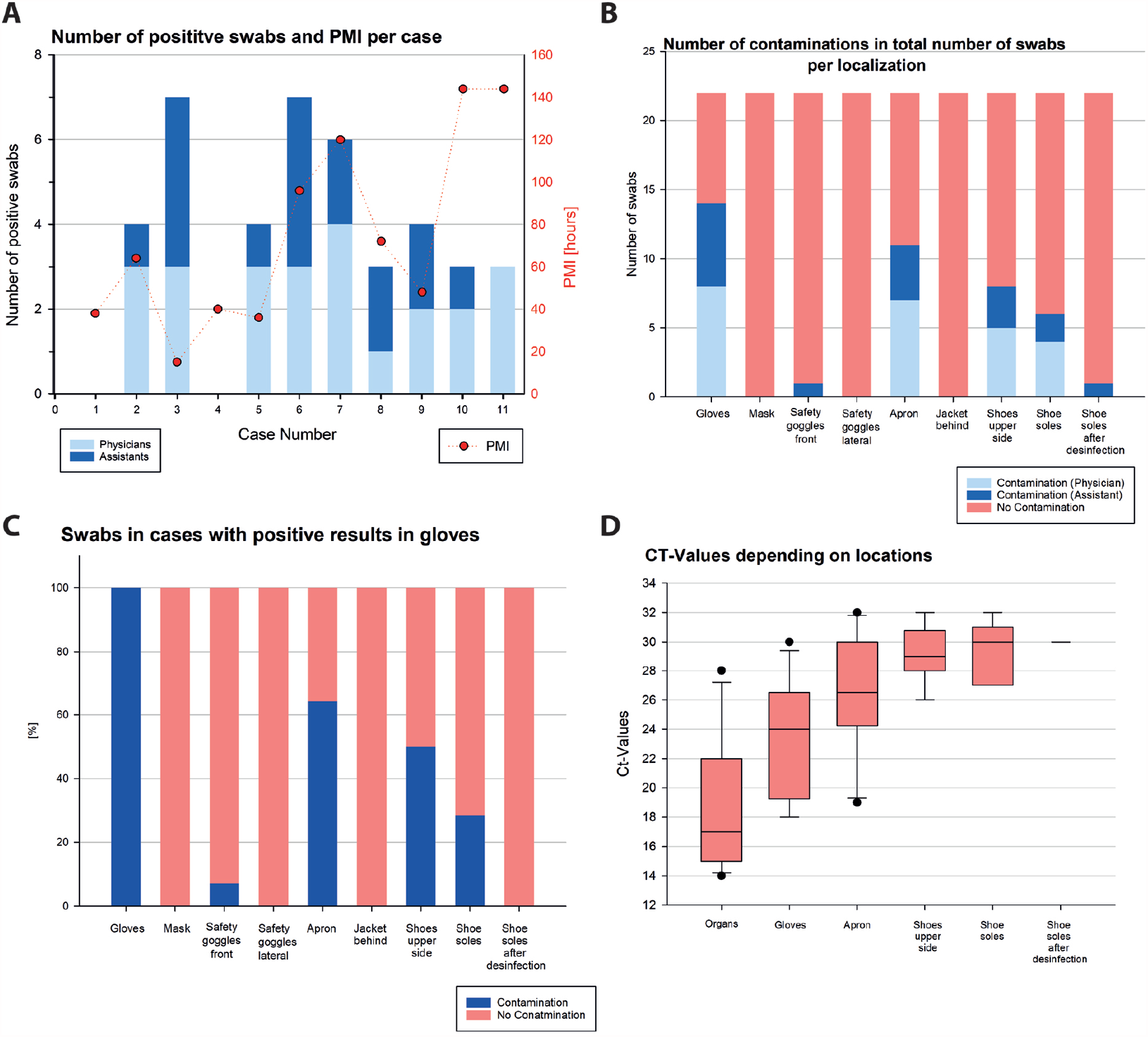
RT-qPCR results from PPE. **a** Number of positive swabs per case divided according to physicians and assistants with corresponding postmortem intervals (PMI). **b** Proportion of positive swabs from physicians and assistants in the different localizations. **c** Results from PPE other than gloves in cases when the gloves were tested positive. **d** Box plots of the Ct-values depending on the localizations.

Contaminations occurring at different locations are shown in Fig. 2B and 2C and Supplementary Fig. 1. Gloves were the most frequently contaminated parts of the PPE (14/22, i.e. 14 SARS-CoV-2 positive samples from 22 glove samples in total, 64%), followed by aprons (11/22, 50%), upper sides of the shoes (8/22, 36%) and shoe soles (6/22, 27%). The front of the safety goggles was positive in 4.5% of the goggle-samples, while lateral sides of safety goggles, FFP masks, and the back of the protective clothing were negative. In the 14 events when gloves were positive, also aprons were positive in 9 events (64%) and the upper sides of the shoes in 7 events (50%) (Fig. 2C). There was a correlation trend between contamination of gloves and aprons (p =0.08). Furthermore, a correlation trend between parallel RNA detection on the apron and the upper sides of the shoes (R = 0.38; p = 0.08) could be observed. The latter was also trend-wise associated with the positivity of the shoe soles (R = 0.39; p = 0.08; Figures 2B and 2C).

A highly significant difference (p < 0.001) was observed between the Ct values of the samples obtained from the lungs or bronchus (median Ct: 17; range: 14 – 28) and from the PPE (median Ct: 28; range: 18 – 32). The viral density decreased highly significantly in a sequel from the lung/bronchus samples (positive controls) via the gloves and the aprons to the shoes (Figure 2D).

### RT-qPCR results of swabs in minimally invasive autopsies

In total, 30 swabs were performed for three autopsies, including one lung control from each autopsy. In one autopsy (MIA-1), the lung control was tested positive for SARS-CoV-2, the lung controls of the other two autopsies (MIA-2 and MIA-3) were negative (Table 1). In MIA-1 viral RNA could be detected from the gloves of the ultrasound-physician, all other swabs were negative. For MIA-2 and MIA-3 all swabs were tested negative for SARS-COV-2 RNA.

### Isolation of infectious virus from full autopsy samples

198 swabs from PPE and 11 swabs from lung/bronchus were taken in parallel with the swabs for RT-qPCR and were stored at -80°C for assessment of infectivity by virus isolation. Virus isolation was performed on the 52 samples from locations where the duplicate swab was tested SARS-CoV-2 RNA positive in AU. Eleven of these samples represented references directly obtained from the lungs/bronchus of the dead bodies. The remaining 41 samples came from the positive PPE locations detected with RT-qPCR before.

Concerning lung/bronchus samples, virus isolation was successful in 4/11 (36%) samples taken at three centers (AU, DR, and HH) (Fig. 3).

**Fig. 3.**
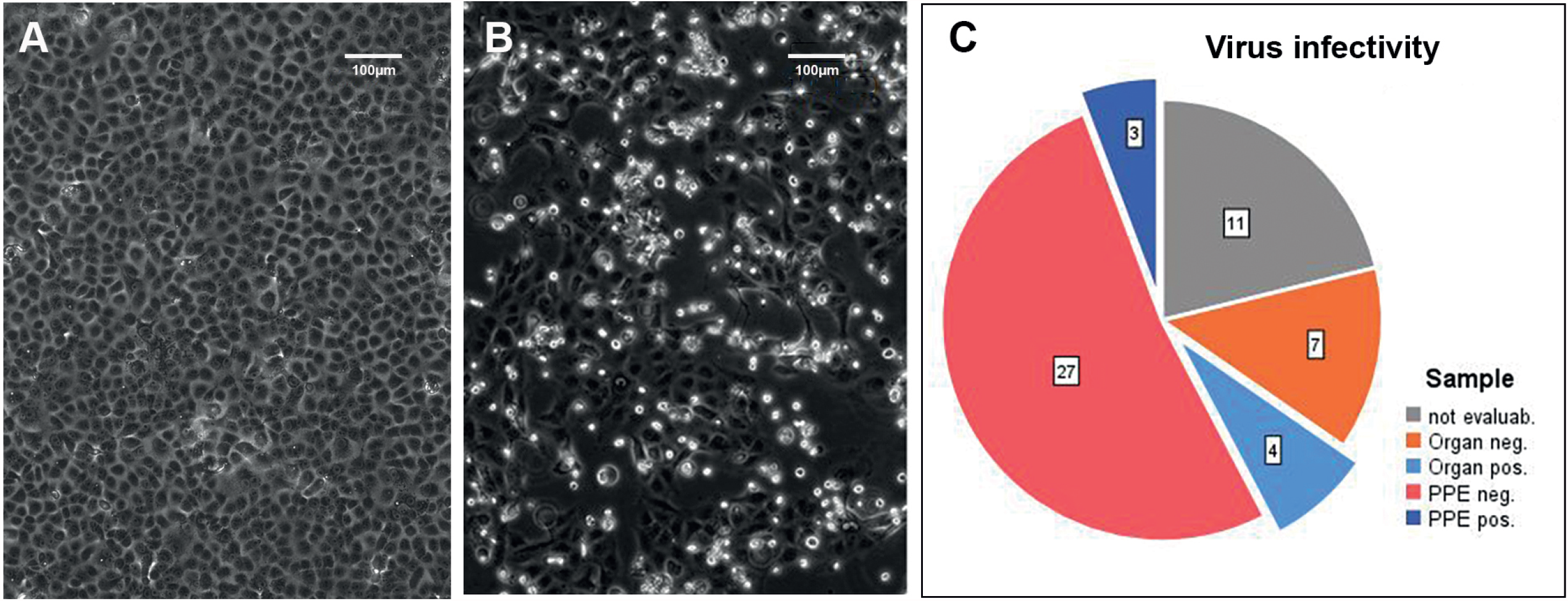
Exemplary representation of the cytopathic effect by SARS-CoV-2 in cell culture. **a** uninfected Vero E6 cells grow to confluence in cell culture, while **b** infected Vero E6 cells show a clear cytopathic effect already 48 hours post infection, characterized by rounding and detachment. **c** Overview of swab samples from organs (lung/bronchus) or PPE positive or negative for successful virus isolation reflecting virus infectivity.

Concerning PPE samples, 11/41 samples used for virus isolation were negative in confirmatory RT-qPCR performed in HH and thus probable RNA degradation was suspected. Hence, they were excluded from the final calculation of infectivity rate. For the sake of completeness, it is mentioned that none of those samples yielded infectious virus.

Out of the remaining 30 PPE samples, isolation of infectious virus was successful in three samples (3/30, 10%). All positive samples came from the gloves (1 assistant-AU, 1 assistant-DR, 1 physician-DR). Thus, 21% (3/14) of the RNA positive glove samples were infectious and samples from three of 11 autopsies (27%) were infectious. In these autopsies, the median time between death and autopsy was 72 hours (range: 15 – 120 hours). The total number of positive swabs per autopsy did not correlate with the PMI (R = 0.049; P = 0.89).

On an autopsy related basis, positive results for virus isolation from positive controls or PPE swabs were obtained in four of six autopsies with a lung/bronchus sample Ct value below 18, suggesting a high viral load, while only one of five cases with lower virus concentration (Ct>21) was infectious (for Ct values, see Table 1). However, this distribution trend did not reach significance (p = 0.242).

## Discussion

This study aimed to evaluate the extent and severity of PPE contamination from physicians and autopsy assistants with SARS-CoV-2 during autopsies of COVID-19 deceased at five German centers under real-world conditions. These five centers represent four different geographic regions (north, south, east, and west), two different medical disciplines (pathology and legal medicine), and two different techniques (conventional complete autopsies and minimally invasive autopsies). Swabs were chosen as the method for generating samples for RT-qPCR and infectivity assessment in cell culture experiments, as used by others recently (10-12).

For full autopsies, only autopsies with positive rapid SARS-CoV-2 diagnostic at the beginning of autopsy were included. Consequently, all of them were SARS-CoV-2 RNA positive in the lungs/bronchus samples used for reference (positive control). Because rapid analysis was not available at MU, this criterion was not fulfilled, and only in one of the three autopsies, the positive control sample (lung biopsy) was tested positive. Thus, the number of evaluable MIA autopsies was only one. The fact that in this one positive case the glove sample was positive showed that even in MIA, PPE contamination could not be completely excluded. However, further investigations with a higher number of autopsies are necessary to elucidate its real extent.

For full autopsies, SARS-CoV-2 RNA contamination of parts of the PPE was found at a high frequency (9/11 autopsies, 82%). In only two out of 11 full autopsies, neither the physician’s nor the assistant’s PPE were contaminated. Pomara et al. reported a considerably lower positivity rate of 15.6% (11). However, they only evaluated face shields. In our study, the front of safety goggles was contaminated only in a single case, resulting in an even lower positivity rate (1/22 goggle samples, 4.5%), compared to Pomara et al., while gloves (64%), aprons (50%), and shoes (upper sides: 36%, soles: 27%) were frequently positive. The distribution of the contaminated PPE parts indicates that intensive mechanical contact is a cause of contamination. As expected, samples from the gloves were tested positive most often, followed by swabs from the apron. Handling the cadavers and the organs makes it difficult to avoid any contact simply because the examiner’s distance and the specimen are naturally minimal in this case. The virus contaminated material, very likely from gloves and aprons, reach the shoes and the floor, from where the shoe soles are also contaminated. This spatial sequence is supported by a stepwise increase in the Ct values, denoting a decreasing viral RNA load (Figure 2D).

Because all samples from the FFP masks, the lateral parts of the safety goggles, and the back of the protective clothing were negative for viral RNA, aerosols are likely not a relevant contamination source in the autopsy setting. However, we cannot exclude the possibility that the viral RNA might be less stable on these surfaces. Nonetheless, direct touch and splash represent the main threats to the transmission of viral material. This direct transmission is likely to be independent of the kind of pathogen. This substantiates the necessity of proper PPE and hygiene measures including waste disposal during and after autopsies.

In concordance with other reports investigating the persistence of viral material on and in people deceased due to COVID-19 ‘(10-12, 17, 18), the PIM of up to 144 hours did not reduce the risk of PPE contamination with SARS-CoV-2 RNA. In our study, we did not aim to evaluate the stability of the virus on PPE over time. Data addressing the topic of SARS-CoV-2 stability have been summarized by Meyerowitz et al. (5).

To assess for infectivity, frozen swabs from all locations previously tested SARS-CoV-2 positive with RT-qPCR were selected for further cultivation, including the reference lung/bronchus samples. Notably, only 41 of these 52 samples tested positive by confirmatory RT-qPCR in HH; therefore, they had to be excluded from calculation of the overall infectivity rate. Retrospectively, it is difficult to identify the reason for this RT-qPCR negativity. RNA degradation during storage and transport could have been an issue. Of note, even though all samples were shipped on dry ice by an experienced courier service, it is remarkable that none of the negative samples came from HH, where the samples could be shipped in-house. Remarkably, the non-evaluable samples were mainly from the shoes (7/11) with high Ct-values. In addition, it is conceivable that sampling the swabs at locations directly next to each other might contribute to these differences, suggesting that the contamination might be more locally concentrated than distributed.

In 3/30 evaluable (i.e. positive in confirmatory RT-qPCR) PPE samples, the isolation of infectious virus was successful. Of note, virus isolation only succeeded from gloves. Besides viral load, which might be highest at the gloves that are in direct contact with the organs, also a more fluid microenvironment on the gloves at the time of sampling may contribute to viral infectivity in contrast to dried out viral material on other PPE items. To our knowledge, this is the first description of isolation of infectious SARS-CoV-2 from PPE in an autopsy setting. Pomara et al. demonstrated SARS-CoV-2 RNA on 15.6% of the face shields, but they did not investigate infectivity (11). Concerning other extracorporal surfaces, Schröder et al., demonstrated viral RNA on six body bags. However, no viable virus was detectable (12).

In the lung/bronchus samples that were chosen as positive control, virus isolation was successful in 4/11 samples (36%). There was a clear trend towards a higher viability rate in cases with low Ct -values under 18 in the lung/bronchus samples, reflecting that higher viral load likely results in higher probability of virus infectivity, as was shown before (19). There was no correlation between successful virus isolation and PMI. Also Plenzig et al. reported the isolation of viable viruses from lungs in two out of four cases independent from PMI (10).

The fact that only the lung/bronchus samples and samples from the gloves were infectious - with a higher infectivity rate in lungs/bronchus - while no infectious virus could be isolated from RNA positive samples from aprons or shoes might indicate a certain instability of SARS-CoV-2 as soon as it is transferred to inanimate surfaces. Haddow et al. investigated SARS-CoV-2 stability on different PPE materials (different face shields, coverall, 50/50 nylon/cotton ripstop fabric) in an experimental setting, and found a PPE material dependent reduction in plaque-forming units over 72 hours (20). For other pathogens, infectivity after transfer to surfaces may be higher or lower, depending on the nature of the pathogen (6, 21).

## Conclusion

The results of this study show a considerable contamination rate of PPE during autopsies of COVID-19 deceased. Contamination even occurred during a minimally invasive approach. Independent of the length of the postmortem interval, SARS-CoV-2 RNA was detectable in 21% of the samples taken from PPE of N = 9 of 11 full autopsies. Gloves (64%), aprons (50%), and shoes (36%) showed the highest frequency of RNA contamination. Infectious virus could be isolated from 21% of the RNA-positive glove samples of N = 3 of 11 full autopsies.

In conclusion, as recommended by several national and international instances, the use of adequate PPE is mandatory because the risk of infection during autopsy is a matter of reality, not a theoretical consideration. Together with hygiene measures, including appropriate waste disposal, they enable the safe performance of COVID-19 autopsies, which are essential for a better understanding of this disease. Also, for future infectious diseases advised selection of appropriate PPE and hygienic measures will provide the basics to carry out autopsies as an important source for new knowledge.

## Data Availability

The datasets used and/or analyzed during the current study are available from the corresponding author on reasonable request.

## Acknowledgments

The authors thank Dr. Susanne Pfefferle, Institute of Microbiology, Virology and Hygiene, University Medical Center Hamburg-Eppendorf, Hamburg, Germany for virus isolation, as well as Nadine Eismann, Christian Beul, Jenny Müller, Marc Britz, and Patrick Kühl for their excellent technical assistance. The authors are also thankful to Oliver Eger for the creation of the illustrations. This study was supported by the German Registry of COVID-19 Autopsies (DeRegCOVID; www.DeRegCOVID.ukaachen.de; funded by the German Federal Ministry of Health, Project No. ZMVI1-2520COR201) and the German Federal Ministry of Education and Research in the framework of the Network of University Medicine (DEFEAT PANDEMIcs, Project No. 01KX2021) as well as the “Bavarian State Ministry for Science, Research and Arts”

## Ethics Approval / Consent to Participate

This study was approved by the ethics commissions of Aachen (EK119/20, EK092/20), Augsburg (BKF No. 2020-18), Dresden (BO-EK-175052020), Hamburg (PV7311) and Munich (20-426). Written consent was obtained from the next of kin to perform the autopsies.

## Author Contribution Statement

DEFEAT PANDEMICs workgroup performed study concept and design, DEFEAT PANDEMICS workgroup, SD and LB performed laboratory analyses and data interpretation, AK and JS-H provided data acquisition and interpretation, JMB and BM provided analysis of data, BM provided statistical analysis, DEFEAT PANDEMIcs workgroup, PB JS-H, SD performed writing of the paper. All authors read and approved the final version of the paper.

## Supplementary Information

**Supplementary figure 4:**
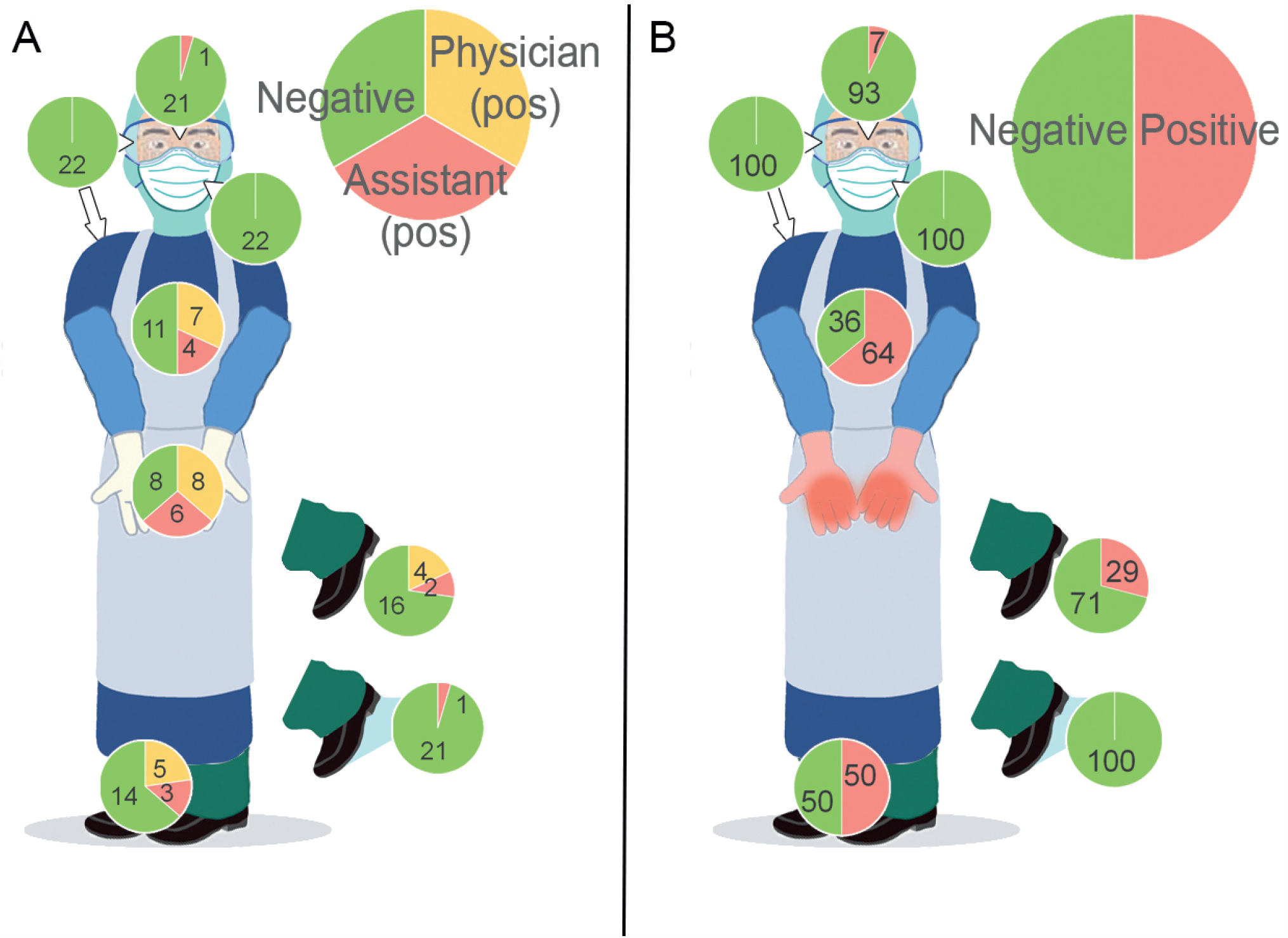
**A** Frequency of contamination of PPE of physicians and assistants at different locations. **B** Results from PPE other than gloves in cases when the gloves were tested positive.

